# A Multi-Laboratory Evaluation of Commercial Monkeypox Molecular Tests

**DOI:** 10.1101/2022.11.27.22282791

**Authors:** Oran Erster, Itzchak Levy, Areej Kabat, Batya Menasheh, Virginia Levy, Hadar Assraf, Roberto Azar, Haim Ben-Zvi, Rita Bridenstein, Olga Bondar, Ayman Fadeela, Ayelet Keren-Naus, Avi Peretz, Diana Roif-Kaminsky, Lolu Saleh, Lisita Schreiber, Orna Schwartz, Pninit Shaked, Nadav Sorek, Merav Strauss, Rachel Steinberg, Orit Treygerman, Simona Zisman-Rozen, Ruth Yshai, Noa Tejman-Yarden, Ella Mendelson, Danit Sofer

## Abstract

In this report, we describe the first national scale multi-laboratory evaluation of commercial quantitative PCR kits for detection of Monkeypox virus (MPXV) DNA. The objective of this study was to assess the performance of two kits by different diagnostic laboratories across Israel. A panel of 10 standardized samples was tested simultaneously using the Novaplex (15 laboratories) and Bio-Speedy (seven laboratories) kits. An in-house assay based on previously published tests was used as reference. Comparison of the results showed high intra-assay consistency between laboratories, with small variations for most samples.

The sensitivity of the two kits was similar to that of the in-house assay, with an analytical detection limit of less than ten copies per reaction. Significant differences were observed, however, in the Cq values and relative fluorescence (RF), between the assays. The RF signal of the in-house and Bio-Speedy assay ranged between 5,000 and 10,000 RFU, while the signal in the Novaplex assay was less than 600 RFU. Due to the kit measurement protocol, the Cq values of the Bio-Speedy kit were 5-7.5 cycles lower than those of the In-house assay. On the contrary, the Cq values of the Novaplex kit were significantly higher than those of the in-house assay, with differences of 3-5 cycles per sample.

Our results suggest that while all assays were similar in their overall sensitivity, direct comparison of Cq values between them may be misleading. Additionally, the low fluorescence obtained with the Novaplex kit may be problematic with marginal or “noisy” samples. Diagnostic laboratories should therefore consider all these aspects when choosing a specific MPX detection assay.

## Introduction

The recent Monkeypox (MPX) outbreak reached a worldwide scale within a few months, including Europe, North and South America, the middle east, Australia and large parts of Asia. This is in addition to Africa, where it has been endemic since its discovery in 1958 (Brown and Leggat 2016). Until October 2022, approximately 70,000 confirmed cases were reported, with actual numbers expected to be significantly higher (Perez Duque et al. 2022). Due to its contagious nature on one hand, and the Prodromal period on the other hand, rapid detection of the virus is important for timely action of proper public health measures (Di Giulio et al. 2004).

Diagnosis of MPX was formerly based on a combination of clinical and epidemiological criteria, and virus isolation (Ladnyj et al. 1972). Later on, diagnosis was made based on electron microscopy, immunological tissue staining, serological tests and, occasionally, on molecular tests (Di Giulio et al. 2004). While virus isolation is the ultimate proof for infection, it cannot be performed in most diagnostic laboratories, it is time-consuming, and cannot be scaled up for high throughput testing. Serological tests can be misleading in some cases, due to the cross-reactivity of Orthopoxviruses, and electron microscopy testing can confirm the presence of Orthopoxvirus, but cannot be performed in most laboratories and cannot identify the species. Until the 1990’s, large populations were vaccinated against smallpox, using vaccinia virus-derived vaccines that generated a cross-protective response. Therefore, such a person would be tested serologically positive for vaccinia, Variola or MPX (Dubois and Slifka 2008). Although serological MPX-specific tests were reported (Dubois and Slifka 2008), such an assay cannot be easily implemented in a large scale testing, and can only be performed in specialized laboratories. This is in addition to the diagnostic limitation associated with the time that takes for development of a detectable seroconversion.

Specific detection of MPX using quantitative PCR (qPCR) is cheaper, faster, and can readily be adjusted for high throughput testing of very large numbers of samples, as was performed during the COVID19 pandemic (Cohen et al. 2022). furthermore, this approach can easily distinguish between different MPX strains (Li *et al*. 2010), which is currently not possible using serological tests. Lastly, qPCR can detect the presence of viral DNA prior to the onset of symptoms, while serological reaction can only be detected 1-2 weeks after exposure. Taken together, these advantages render molecular testing the preferred choice for detecting MPX infection. However, due to the fact that the disease was geographically limited until the present outbreak, availability of commercial MPX detection kits was very limited, until recently.

MPX was first reported in Israel during 2018, when an Israeli resident returned from Nigeria following exposure to infected rodents (Erez et al. 2018). This patient was isolated and recovered without any known subsequent infections. On 21^st^ of May 2022, the first MPX-confirmed case was reported in Israel, and by October 2022, approximately 250 confirmed cases were reported (Mathieu E et al. https://ourworldindata.org/monkeypox). In order to enable a country-wide diagnostic capacity, to respond to the increased circulation of MPX, the Israel Ministry of Health initiated a national-scale qualification procedure for MPX diagnosis, in which participating laboratories performed a qPCR test on a standardized panel of samples. The results were analyzed by the Israel Central Virology Laboratory and are presented in this report.

## Methods

### Sample preparation

The samples used for the study were prepared from the remaining of DNA extractions of Monkeypox clinical samples that were obtained from patients during routine diagnostics. Samples were used for this study under the institutional committee of the “SHEBA ethical board” under protocol number 9481-22-SMC. Five individual samples were pooled together and diluted to generate a set (panel) of eight pools in different viral DNA concentrations. Additional two samples of Varicella Zoster virus (VZV) and Orf virus (OrfV) were included in the sample set as Monkeypox-negative samples. Each pool dilution was aliquoted and stored in -80°C.

### CVL MPXV qPCR assay

The reference test that was used in this study was based on the reactions developed by Li *et al*. (2010) for general detection of MPXV and specific detection of Clade II (formerly West Africa) MPXV strain. The primers and probes described by Li *et al*. were incorporated into a single assay, together with primers and probe targeting human RNAseP as an internal control (https://www.cdc.gov/coronavirus/2019-ncov/lab/rt-pcr-panel-primer-probes.html). The forward primer of the GE reaction was modified from the original design, so that it contained either A or G in position 6 from the 5’p, to complement recently sequenced samples. The sequence of this primer was therefore 5’-GGAAA**R**TGTAAAGACAACGAATAC-3’, where R (A or G) replaces the original A. The multiplex assay was tested for its sensitivity and specificity, using serial dilutions of a clinical sample, and clinical non-MPXV samples of other pathogens. Table 1 details the components that were assembled in the reaction mix.

**Table 1.** Average and standard deviation Cq values of the standard MPXV samples panel obtained with the CVL assay based on the reactions described by Li *et al*. (2010).

| CVL MPX assay |  |  |  |  |  |  |  |
| --- | --- | --- | --- | --- | --- | --- | --- |
| AVG |  |  |  | STDEV |  |  |  |
| Sample | GE | WA | RNASEP | Sample | GE | WA | RNASEP |
| MPX 1 | 27.4 | 28.4 | 33.6 | MPX 1 | 1.29 | 0.78 | 1.7 |
| MPX 2 | 34.9 | 35.7 | 37.3 | MPX 2 | 0.62 | 0.53 | 1.5 |
| MPX 3 | 32.1 | 32.6 | 38.2 | MPX 3 | 1.36 | 1.49 | 1.5 |
| MPX 4 | 21.9 | 23.0 | 28.0 | MPX 4 | 0.59 | 0.64 | 1.3 |
| MPX 5 | 29.4 | 30.5 | 36.2 | MPX 5 | 0.93 | 0.70 | 1.6 |
| MPX 6 |  |  | 34.7 | MPX 6 |  |  |  |
| MPX 7 | 35.4 | 36.2 | 34.5 | MPX 7 | 0.52 | 0.78 | 1.2 |
| MPX 8 | 25.7 | 26.8 | 32.9 | MPX 8 | 0.82 | 0.76 | 1.6 |
| MPX 9 | 29.6 | 30.5 | 36.1 | MPX 9 | 0.55 | 0.77 | 0.4 |
| MPX 10 |  |  | 31.9 | MPX 10 |  |  | 0.41 |

The following run protocol was used

(1) 45°C for 5 min., (2) 2 95°C for 4 min., 40 X[(3) 95°C for 4 min., (4) 58°C for 10 sec., (5) 60°C for 0:10 + Plate Read]. Graphical representation of the protocol is shown on **supplementary Figure S1**.

### Seegene Novaplex MPX kit

The Seegene Novaplex kit Cat. no. R-PX10325Z for detection of MPXV DNA (https://www.seegene.com/assays/novaplex_mpxv_assay) was used according to the manufacturer’s instructions. Importantly, the kit internal control was spiked into the samples before setting the reaction. Graphical representation of the amplification program is shown on **supplementary Figure S1**.

### Bio-Speedy Monkeypox kit

The Bio-Speedy Monkeypox virus qPCR kit (Bioeksne.com.tr, Cat. No. BS-MPV-25) was prepared and used according to the manufacturer’s instructions (bioeksen.com.tr/en/Monkeypox-1). Graphical representation of the amplification program is shown on **supplementary Figure S1**.

### Statistical analysis

The average and standard deviation were calculated for each sample.

## Results

### Evaluation of the CVL CDC-based In-house assay

The sensitivity of the “general” (GE) and “West African” (WA) reactions was determined by Li *et al*. (2010). Since we combined three reactions together, we sought to ensure that the specificity was retained and that there were no non-specific cross-reactions of the components, resulting in a false signal. The analytical limit of detection (LOD) for the multiplex assay was therefore determined using a PCR product corresponding to the TNF receptor gene region. Serial dilutions of the PCR product were tested, as shown in **supplementary Figure S2**, demonstrating a sensitivity of 2 copies/µl for both MPXV reactions. The numerical values of the analysis are detailed in **supplementary Table S1**. Specificity was evaluated by testing samples of Zoster virus (VZV) Measles virus (MeV) and Orf virus, and were all negative (**Table S1**). This assay was then integrated into the diagnostic routine of the Israel Central Virology Laboratory (CVL) and is currently used to diagnose suspected MPX samples. This assay was used to set the reference values for the study with the samples panel. The standard sample panel was tested six times on different days, and the average values and standard deviation obtained are detailed in **Table 2**. As samples with MPXV Cq values greater than 31 contain diluted samples, their respective RNAse P were high, leading to a larger standard deviation (STDEV). For the MPX reactions, the STDEV ranged between 0.36 to 1.36 cycles (**Table 2**).

**Table 2.** Average Cq values and standard deviation of the samples panel tested with the Novaplex kit in fifteen diagnostic laboratories. In the right column, the Cq value differences between the average CVL In-house assay and the average Novaplex assay are detailed.

| Sample | Average Cq | STDEV |  | Cq<sub>NPVPLX</sub>-Cq<sub>CVL</sub> |
| --- | --- | --- | --- | --- |
| MPX 1 | 32.8 | 1.76 |  | 4.0 |
| MPX 2 | 40.4 | 0.90 |  | 4.6 |
| MPX 3 | 37.5 | 1.02 |  | 4.2 |
| MPX 4 | 26.5 | 1.15 |  | 3.3 |
| MPX 5 | 34.3 | 2.69 |  | 3.7 |
| MPX 6 |  |  |  |  |
| MPX 7 | 41.3 | 1.07 |  | 4.7 |
| MPX 8 | 29.8 | 1.33 |  | 2.7 |
| MPX 9 | 35.4 | 3.02 |  | 5.2 |
| MPX 10 |  |  |  |  |

### Comparative evaluation of the Novaplex kit

In order to evaluate the performance of the Seegene Novaplex MPX kit, the standardized MPXV samples panel was tested in parallel by 15 diagnostic laboratories across Israel. The list of participating laboratories is detailed in **supplementary Table S2**. The panel samples were randomly labeled 1-10, without any indication of the expected result. Each laboratory performed the test in a duplicate and the results were analyzed. All laboratories identified 7 out of the 8 positive samples, and seven identified marginal sample MPX7 as positive. The results of each laboratory with respect to each other and to the CVL In-house assay are depicted in **Figure 1A** and the average values of each sample are shown in **Table 2**. The values detailed under the “CVL In-house” test were obtained from the WA reaction, which is more stringent than the GE reaction (Li *et al*. 2010). All laboratories correctly identified the VZV and Orf virus samples as negative for MPXV. Positive samples MPX1, 2, 3, 5, 8 and 9 were identified as positive by all laboratories (**Figure 1A and Supplementary Table S3**). Sample MPX7, which gave a Cq value similar to that of MPX2 in the CVL In-house assay, was identified as positive by 8 of the 15 laboratories. Within these eight laboratories, in three it was identified in one of two repeats, as detailed in **Supplementary table S3**. The internal control of the kit was spiked into the samples and gave Cq values well below 45, indicating that the samples were not degraded or inhibited.

**Figure 1.**
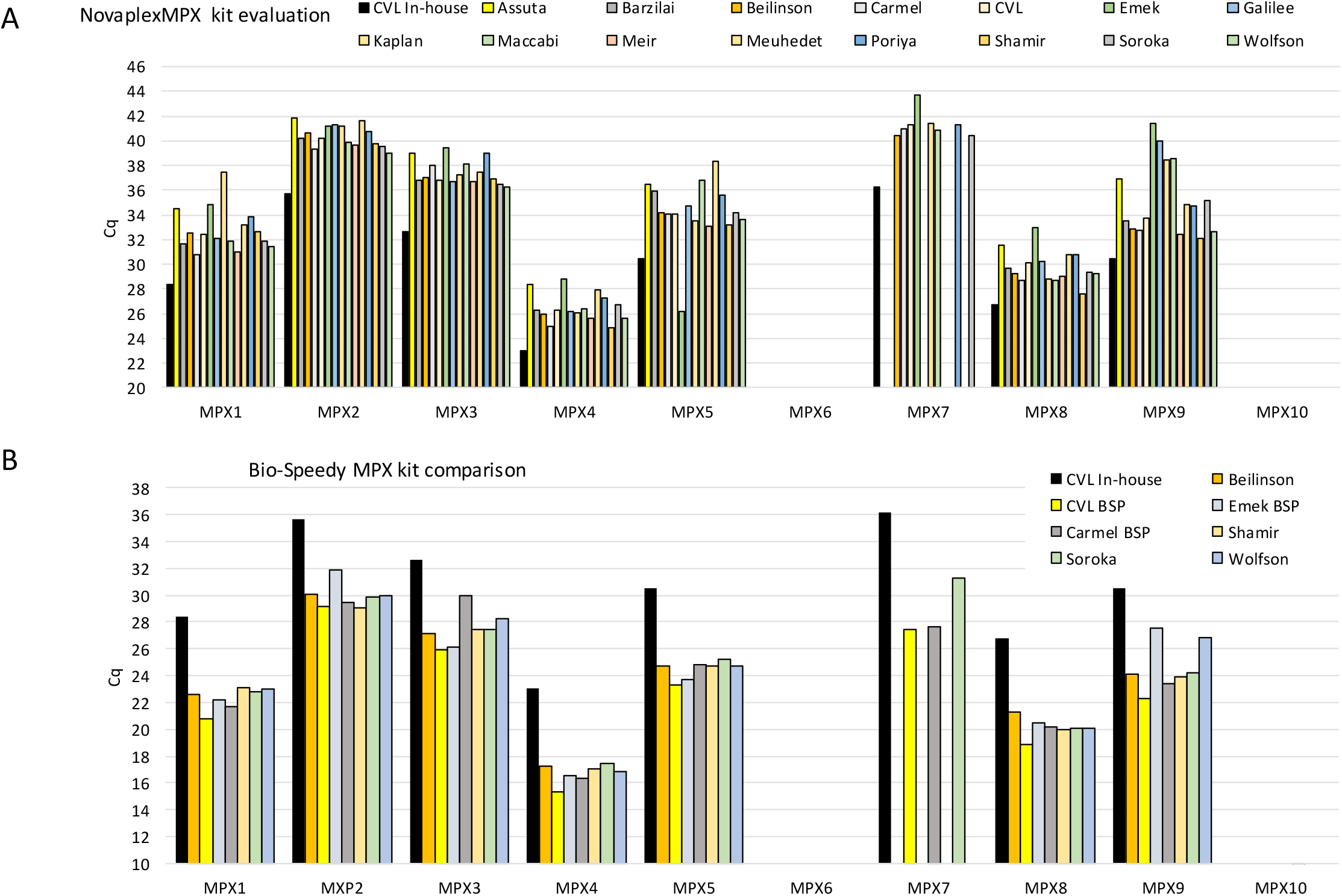
Average Cq values obtained from each participating laboratory, of each MPX panel sample. The average value represents at least two separate repeats performed in each laboratory. (A) Cq values obtained using the Novaplex kit. (B) Cq values obtained with the Bio-Speedy kit.

Comparison of the Cq values obtained with the Novaplex kit from different laboratories, showed a varying degree of deviation (**Table 2, right column**). Samples 1-4 and 8 had a standard deviation of 0.9-2.75 cycles, while sample 5 had a deviation of 2.7 and sample 9 had a deviation of 3 cycles. The marginal sample MPX 7, was identified as positive in eight out of the 15 laboratories, with an average value of 40.5 and a STDEV of 1.1 among the repeats.

Notably, all results obtained by the Novaplex kit, had higher Cq values than those obtained with the CVL assay. The Cq differences of the average values obtained for each positive sample, ranged between 2.7 and 5.2 cycles (**Table 2**).

### Comparative evaluation of the Bio-Speedy kit

In order to evaluate the performance of the Bioeksen Bio-Speedy Monkeypox (MPX) kit (https://www.bioeksen.com.tr/en/monkeypox-1), seven laboratories tested the same sample panel with this kit. The sensitivity and uniformity of the kit performance was similar to that of the Novaplex kit, with all runs identifying seven of the eight positive samples, and sample MPX7, identified in three of the seven labs (**Figure 1B, Table 3**). The variation between the results ranged between 0.7 and 2.1 cycles (**Table 3**). The numerical values obtained by each laboratory are detailed in **supplementary Table S4**). Since the original samples used for the study were highly diluted, the RNAse P control reaction was either very weak or negative for most samples and was therefore not compared here.

**Table 3.** Average Cq values and standard deviation of the samples panel tested with the Bio-speedy kit in seven diagnostic laboratories. The right column shows the Cq value differences between the average CVL In-house assay and the average Bio-speedy assay are detailed, resulting from the delayed measurement in the Bio-speedy protocol.

| Sample | BSP average | BSP STDEV | Cq<sub>CVL</sub>-Cq<sub>BSP</sub> |
| --- | --- | --- | --- |
| MPX1 | 22.3 | 0.8 | 6.1 |
| MPX2 | 29.9 | 1.0 | 5.8 |
| MPX3 | 27.5 | 1.4 | 5.1 |
| MPX4 | 16.7 | 0.7 | 6.3 |
| MPX5 | 24.4 | 0.7 | 6.1 |
| MPX6 |  |  |  |
| MPX7 | 28.8 | 2.1 | 7.4 |
| MPX8 | 20.1 | 0.7 | 6.6 |
| MPX9 | 24.6 | 1.9 | 5.9 |
| MPX10 |  |  |  |

The Cq values obtained with the Bio-Speedy kit were consistently lower than those obtained with the In-house assay, due to the different test protocols. The fluorescence reading starts on the first amplification cycle in the CVL In-house and Novaplex tests, while in the Bio-Speedy test, reading starts after 11 touchdown cycles (**Supplementary Figure S1, Bio-speedy protocol**). This resulted in a shift of 5.1-7.4 Cq values between the two assays **(Table 3, right column**).

Comparison of the relative fluorescent signal generated by the different reactions showed marked differences between the Novaplex assay and the other two assays. **Figure 2** shows representative amplification curves and RFU values generated from two representative samples, MPX4 and MPX9. These samples gave RFU values of 6,000-10,000 units in the CVL and Bio-Speedy assays (**Figure 2A, B**). For the Novaplex assay, both the curves generated directly by the Bio-Rad CFX software and the curves generated by the Seegene Viewer software, gave RFU values of 200-550 units (**Figure 2C, D**).

**Figure 2.**
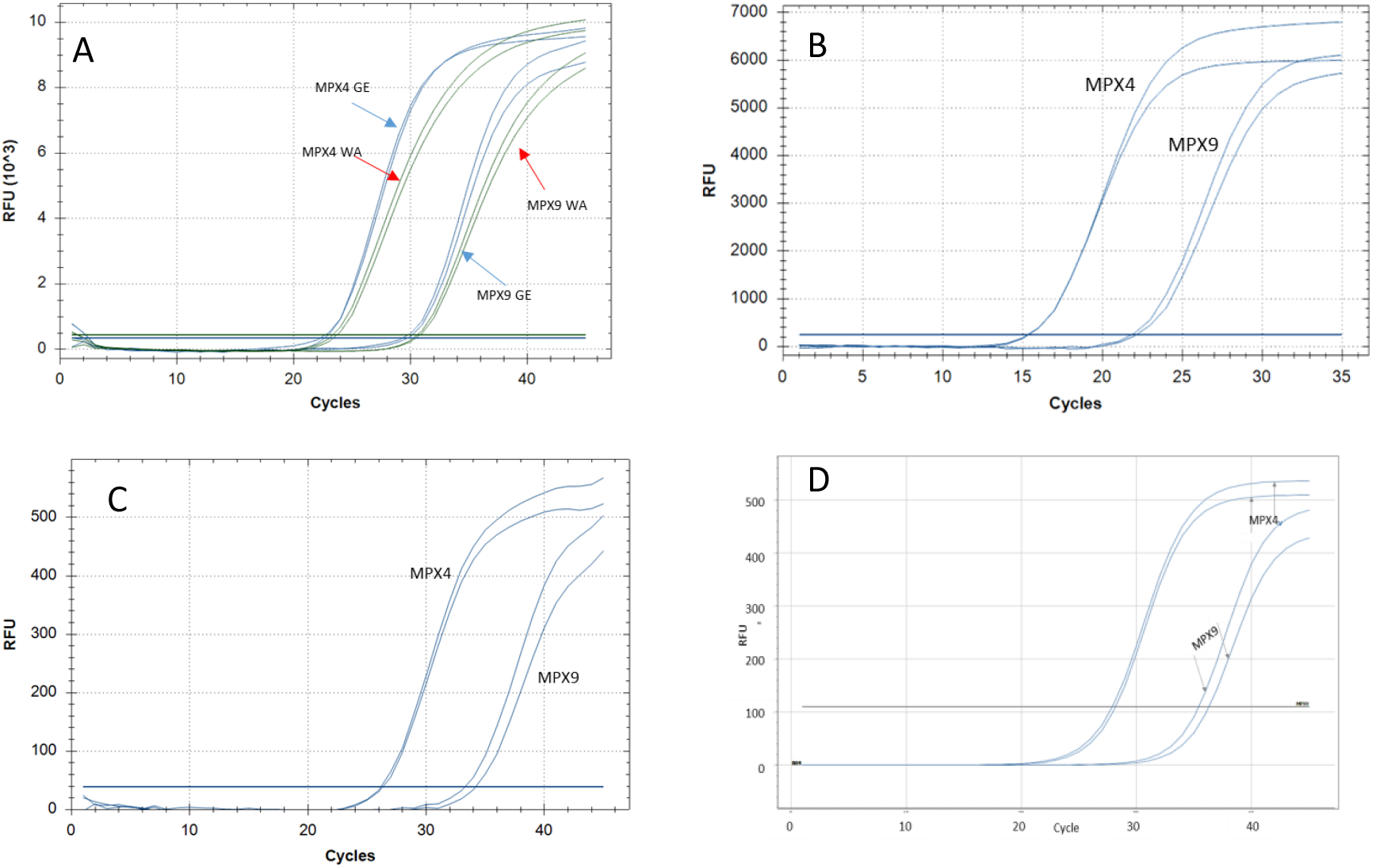
Amplification curves of the CVL assay and the Novaplex assay. Duplicate curves of representative panel samples MPX4 and MPX9 are shown, as obtained with the CVL In-house assay (A), the Bio-Speedy assay (B), the Novaplex raw analysis (C), and the Novaplex accompanying software (Seegene viewer) analysis (D). RFU – Relative fluorescence units.

## Discussion

Although Monkeypox (MPX) was discovered more than 60 years ago, and the first human case was diagnosed more than 50 years ago (Ladnyj et al. 1972), molecular commercial detection kits were not widely available until the current multinational outbreak. The MPX outbreak in the USA during 2003 prompted the development of several molecular assays, using somewhat different technologies with varying specificities towards Orthopox species and Monkeypox in particular (Neubauer et al. 1998, Kulesh et al. 2004, Shchelkunov et al. 2004, Li et al. 2006, Maksyutov et al. 2016). In order to facilitate high throughput testing in a short time and affordable cost, we set to establish a quantitative PCR assay that can identify all MPXV strains, and indicated whether a positive sample belongs to the WA strain or not.

In order to perform a robust and informative assay, we combined the general (GE) and WA clade-specific (WA) reactions described by Li *et al*. (2010) with an internal control reaction and demonstrated that the two MPX reactions are sensitive and have very similar kinetics. A recent pre-print publication (Wu et al. 2022) suggested that changes in primers sequences developed by Li *et al*. should be made, to render them more suitable to detect the currently circulating MPX clade. In our reaction design, we addressed this by altering the sequence of the MPX general (GE) reaction forward primer, already in May 2022, based on genomic analysis of currently circulating MPX isolates. The WA reaction components showed perfect match to sequences of circulating MPX samples and were therefore not altered (Erster *et al*., manuscript in preparation). The assay’s sensitivity was consistent with that described by Li *et al*., indicating that the combined multiplex retained the reactions’ sensitivity. Previous reports of quantitative PCR assays reported similar sensitivity of 5-100 copies per reaction, with very high specificity (Li et al. 2006, Maksyutov et al. 2016). Based on these reports, this assay was used as reference in this study. Upon the onset of the current MPX outbreak, a rapid release of commercial kits was employed, to meet the urgent demand. To facilitate a national scale capability for MPX diagnosis, two commercial assays that include an internal control reaction and can therefore be used as first-line test, were evaluated.

Extensive experience gained during the COVID19 pandemic suggested that while in some cases, different molecular kits showed very similar performance (Dundon et al. 2021), several studies reported on significant differences between different test procedures (Malecki et al.2021, Freire-Paspuel et al. 2021, Wang et al. 2021). Our study focused on evaluating the reproducibility of the two kits by simultaneous testing in different laboratories, but also on comparing the commercial kits and the CVL in-house assay. The Comparison showed high reproducibility for most samples, with acceptable standard deviation values, as shown in **Figure 1** and detailed in **Table 2**. Weak samples resulted in higher SD values, as expected when the reproducibility of the assay declines. The uniformity of the results obtained by 15 laboratories (Novaplex kit) and seven laboratories (Bio-Speedy kit) indicated that the products are sufficiently standardized, with a decline in reproducibility when testing marginal samples (Cq values higher than 40 in the Novaplex kit). The relatively high variability in the results of sample MPX9 is unaccounted for, since all samples originated from one pool of five clinical samples, inhibitory effects or other factors that affect the results should be similar in all samples. As the results of samples with similar or lower DNA concentrations were not as variable, this cannot be explained by differences in the kit quality. This variability was evident both in the Novaplex and Bio-Speedy kits.

Both assays showed a similar limit of detection, which was comparable to that of the CVL in-house assay. The numerical Cq values, however, were significantly different, as was previously reported for SARS-COV-2 kits (Malecki et al. 2021). While the low values obtained with the Bio-Speedy kit resulted from the fluorescence measurement protocol, the values obtained with the Novaplex assay cannot be explained by the amplification protocol and are probably due to the proprietary Novaplex components and the reaction kinetics. These differences suggest that direct comparison between results of different assays cannot be performed and the “Cq shift” should be taken into consideration, when analyzing results obtained with different assays. Importantly, all three assays correctly detected weak samples (marginal sample MPX7 was detected in some of the repeats), so the Cq differences did not significantly affect the performance in terms of sensitivity.

In addition to the consistent shift in Cq values, the relative fluorescence (RFU values) varied significantly, between the in-house and Bio-Speedy assays, and the Novaplex assay, where the RFU values were between 10 and 20-fold lower. While the RFU value by itself does not usually affect the test result, a combination of low fluorescent signal and background “noise” can lead to misinterpretation of marginal results. The Novaplex kit accompanying software (Seegene Viewer for MPX) is somewhat helpful in improving the test results, but cannot completely overcome the potential problems associated with the low fluorescence.

This study has several limitation related to the small number of tested samples (eight positive and two negative), and to the small number of repeats performed by each laboratory. Since the amount of standardized sample material and the availability of the kits were limited, multiple repeats that could determine the limit of detection more conclusively for each lab, were not performed. However, since the results were uniform for most samples, in and between labs, we believe that the data obtained herein are sufficiently significant to support the study’s conclusions. Another limitation is the small number of clinical samples that was used; in order to standardize the evaluation, a pool of five relatively concentrated samples was used as a starting point. This somewhat limits the ability to assess the robustness of the assays, as different samples may contain different compositions of inhibitors that may affect the test results.

In conclusion, both kits can be used for laboratory diagnosis of MPXV, providing that they are used according to the manufacturer’s instructions. However, the weak fluorescent signal and delayed Cq values obtained using the Novaplex kit require attention when testing suspected samples that give marginally positive results. Additionally, comparison of the results obtained by different assays require proper adjustments that take into account the different measurement protocols. It is expected that if the MPX outbreak will persist, existing products will be improved and additional products will become available, thereby improving the diagnostic capacity of Monkeypox infections. It is therefore advisable for diagnostic laboratories to consider the aspects addressed herein, when choosing which assay to use for MPX detection.

To our knowledge, this is the first national scale multi-laboratory comparative study of Monkeypox commercial detection kits.

## Data Availability

All data produced in the present study are available upon reasonable request to the authors.

## Supplementary material

**Supplementary Table S1.**
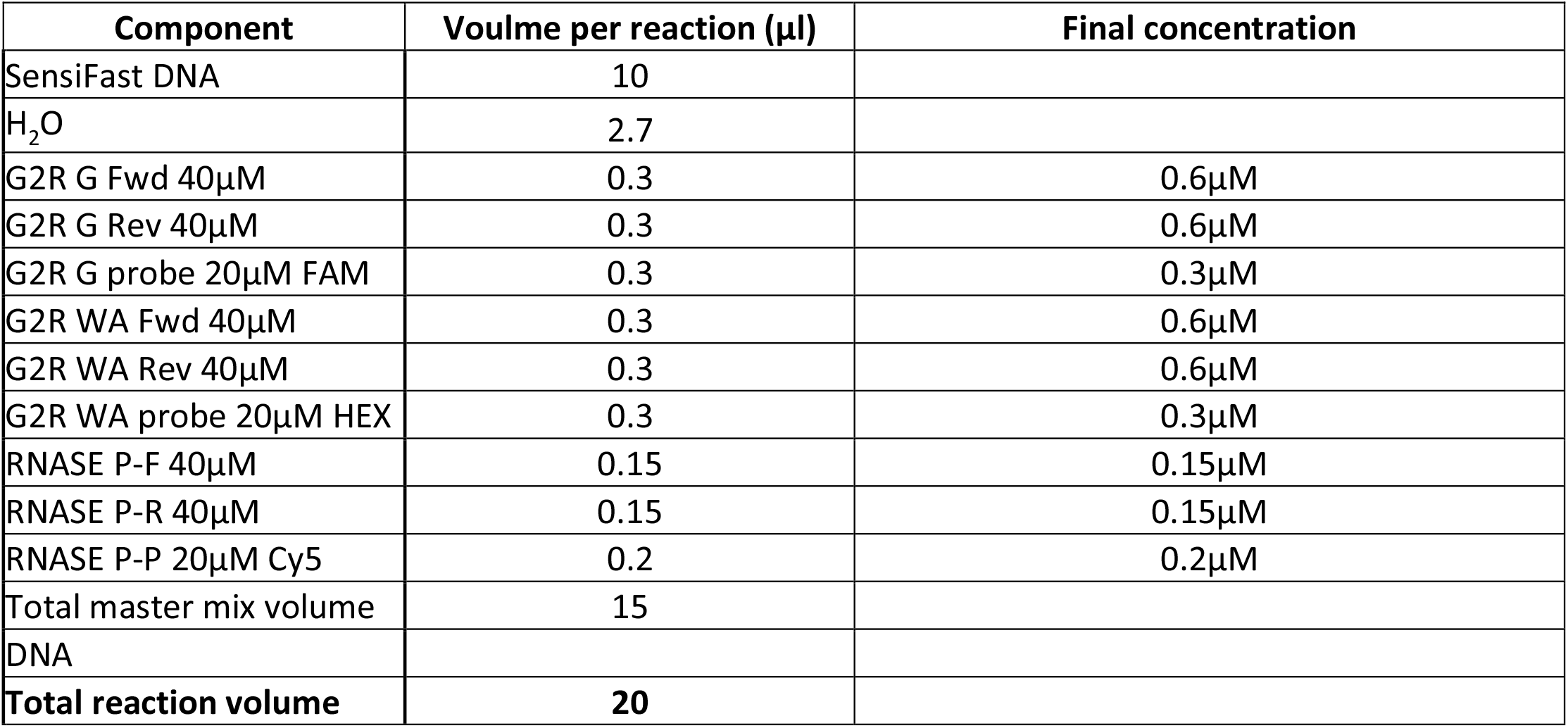
Components of the multiplex CVL MPX assay.

| Component | Volume per reaction (μl) | Final concentration |
| --- | --- | --- |
| SensiFast DNA | 10 |  |
| H<sub>2</sub>O | 2.7 |  |
| G2R G Fwd 40μM | 0.3 | 0.6μM |
| G2R G Rev 40μM | 0.3 | 0.6μM |
| G2R G probe 20μM FAM | 0.3 | 0.3μM |
| G2R WA Fwd 40μM | 0.3 | 0.6μM |
| G2R WA Rev 40μM | 0.3 | 0.6μM |
| G2R WA probe 20μM HEX | 0.3 | 0.3μM |
| RNASE P-F 40μM | 0.15 | 0.15μM |
| RNASE P-R 40μM | 0.15 | 0.15μM |
| RNASE P-P 20μM Cy5 | 0.2 | 0.2μM |
| Total master mix volume | 15 |  |
| DNA |  |  |
| **Total reaction volume** | **20** |  |

**Supplementary Table S2.** List of participating laboratories and their institutional affiliation.

| **Laboratory institutional affiliation** | **Lab abbreviation** |
| --- | --- |
| Assuta Ashdod University hospital | Assuta |
| Barzilai Medical Center | Barzilai |
| Beilinson-Rabin Medical Center | Beilinson |
| Carmel Medical Center | Carmel |
| Israel Central Virology Laboratory | CVL |
| Galilee Medical Center | Galilee |
| Emek Medical Center in Afula | Haemek |
| Kaplan Medical Center | Kaplan |
| Maccabi Helathcare Central Laboratory | Maccabi |
| Meir Medical Center | Meir |
| United ("Me'uhedet") Helathcare Central Laboratory | Me'uhedet |
| Baruch Padeh Medical Center AKA Poriya Medical Center | Poriya |
| Shamir Medical Center, formerly Assaf Harofeh Medical Center | Shamir |
| Soroka Medical Center | Soroka |
| Wolfson Medical Center | Wolfson |

**Supplementary Table S3.** Novaplex comparative test. Average Cq values obtained from fifteen diagnostic laboratories for each panel sample using the Novaplex kit. All laboratories tested the same samples using the same protocol.

| Test | Lab\Sample | MPX1 | MPX2 | MPX3 | MPX4 | MPX5 | MPX6 | MPX7 | MPX8 | MPX9 | MPX10 |
| --- | --- | --- | --- | --- | --- | --- | --- | --- | --- | --- | --- |
| CVL In-house | CVL In-house | 28.9 | 35.8 | 33.2 | 23.2 | 30.6 |  | 36.6 | 27.1 | 30.2 |  |
| Novaplex | Assuta | 34.5 | 41.9 | 39.1 | 28.4 | 36.5 |  |  | 31.6 | 37.0 |  |
| Novaplex | Barzilai | 31.7 | 40.2 | 36.8 | 26.3 | 36.0 |  |  | 29.7 | 33.5 |  |
| Novaplex | Beilinson | 32.6 | 40.7 | 37.0 | 26.0 | 34.2 |  | 40.4 | 29.3 | 32.8 |  |
| Novaplex | Carmel | 30.8 | 39.3 | 38.1 | 25.0 | 34.1 |  | 41.0 | 28.7 | 32.7 |  |
| Novaplex | CVL | 32.4 | 40.2 | 36.8 | 26.3 | 34.1 |  | 41.3 | 30.1 | 33.7 |  |
| Novaplex | Emek | 34.8 | 41.3 | 39.4 | 28.8 | 26.2 |  | 43.8 | 33.0 | 41.5 |  |
| Novaplex | Galilee | 32.1 | 41.3 | 36.7 | 26.2 | 34.7 |  |  | 30.2 | 40.0 |  |
| Novaplex | Kaplan | 37.5 | 41.2 | 37.2 | 26.1 | 33.5 |  | 41.4 | 28.9 | 38.5 |  |
| Novaplex | Maccabi | 31.9 | 39.9 | 38.1 | 26.4 | 36.9 |  | 40.8 | 28.7 | 38.6 |  |
| Novaplex | Meir | 31.0 | 39.6 | 36.7 | 25.6 | 33.1 |  |  | 29.0 | 32.5 |  |
| Novaplex | Me'uhedet | 33.2 | 41.6 | 37.5 | 28.0 | 38.4 |  |  | 30.7 | 34.8 |  |
| Novaplex | Poriya | 33.8 | 40.8 | 39.0 | 27.2 | 35.6 |  | 41.3 | 30.8 | 34.8 |  |
| Novaplex | Shamir | 32.7 | 39.7 | 37.0 | 24.9 | 33.2 |  |  | 27.6 | 32.1 |  |
| Novaplex | Soroka | 31.9 | 39.5 | 36.5 | 26.7 | 34.2 |  | 40.4 | 29.4 | 35.2 |  |
| Novaplex | Wolfson | 31.5 | 39.0 | 36.3 | 25.6 | 33.7 |  |  | 29.2 | 32.6 |  |

**Supplementary Table S4.** Bio-speedy comparative test. Average Cq values obtained from seven diagnostic laboratories for each panel sample using the Bio-speedy (BSP) kit. All laboratories tested the same samples using the same protocol.

| Test | Lab\Sample | MPX1 | MPX2 | MPX3 | MPX4 | MPX5 | MPX6 | MPX7 | MPX8 | MPX9 | MPX10 |
| --- | --- | --- | --- | --- | --- | --- | --- | --- | --- | --- | --- |
| CVL In-house | CVL In-house | 27.4 | 34.9 | 32.1 | 21.9 | 29.4 |  | 35.4 | 25.7 | 29.6 |  |
| BSP | Beilinson | 22.6 | 30.1 | 27.2 | 17.3 | 24.7 |  |  | 21.2 | 24.1 |  |
| BSP | CVL | 20.8 | 29.2 | 26.0 | 15.4 | 23.3 |  | 27.4 | 18.9 | 22.3 |  |
| BSP | Emek | 22.2 | 31.9 | 26.2 | 16.6 | 23.7 |  |  | 20.5 | 27.5 |  |
| BSP | Carmel | 21.7 | 29.4 | 30.0 | 16.4 | 24.8 |  | 27.7 | 20.1 | 23.4 |  |
| BSP | Shamir | 23.1 | 29.1 | 27.4 | 17.1 | 24.7 |  |  | 19.9 | 23.9 |  |
| BSP | Soroka | 22.8 | 29.9 | 27.5 | 17.4 | 25.2 |  | 31.3 | 20.1 | 24.2 |  |
| BSP | Wolfson | 23.0 | 29.9 | 28.2 | 16.8 | 24.7 |  |  | 20.1 | 26.8 |  |

**Supplementary Figure S1.**
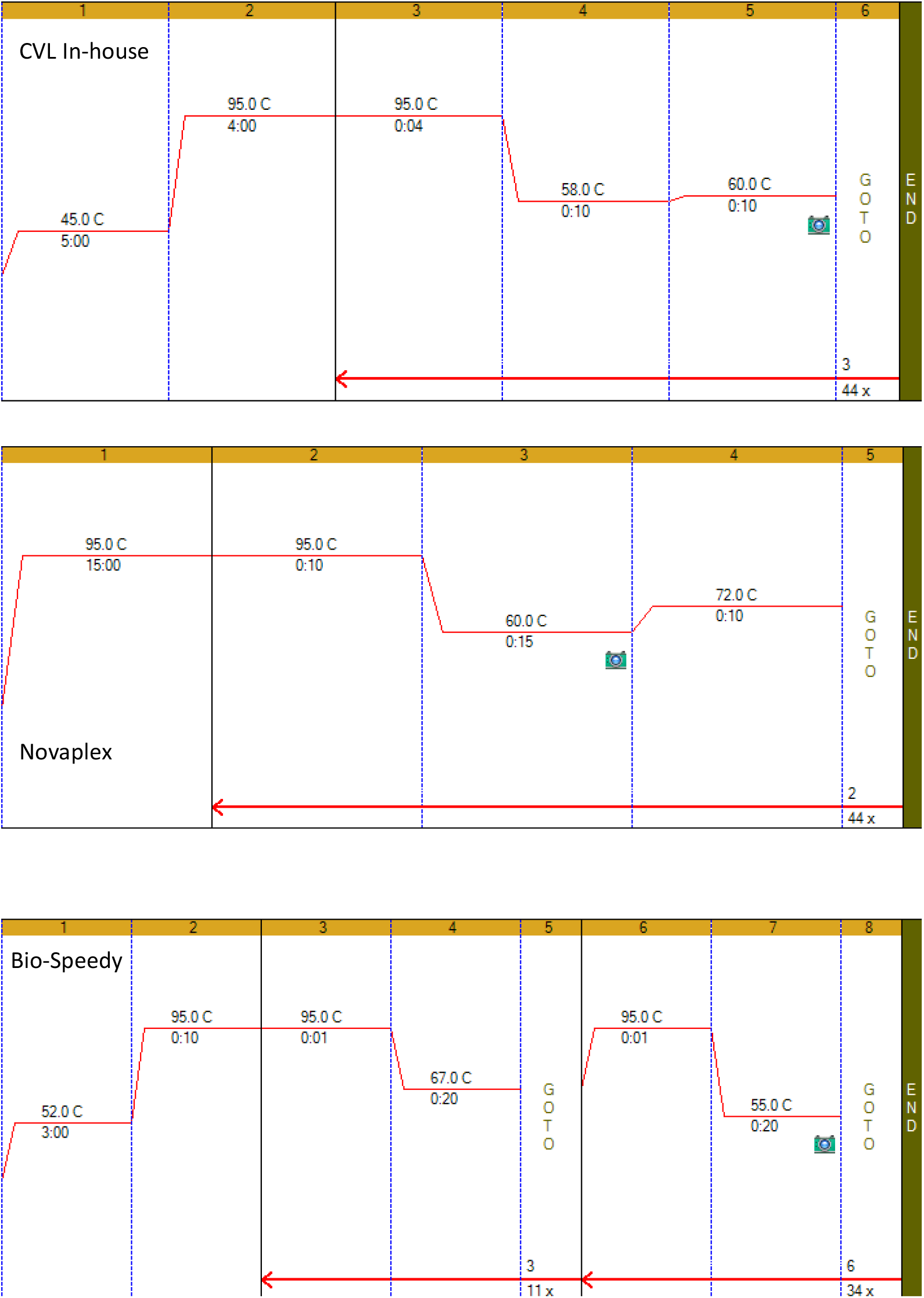
Graphical representation of the cycling protocol of each assay. While fluorescent recording starts on the first amplification cycle in the CVL and Novaplex protocols (top and central panels), in the Bio-Speedy protocol (bottom panel) it starts after 11 cycles.

**Supplementary Figure S2.**
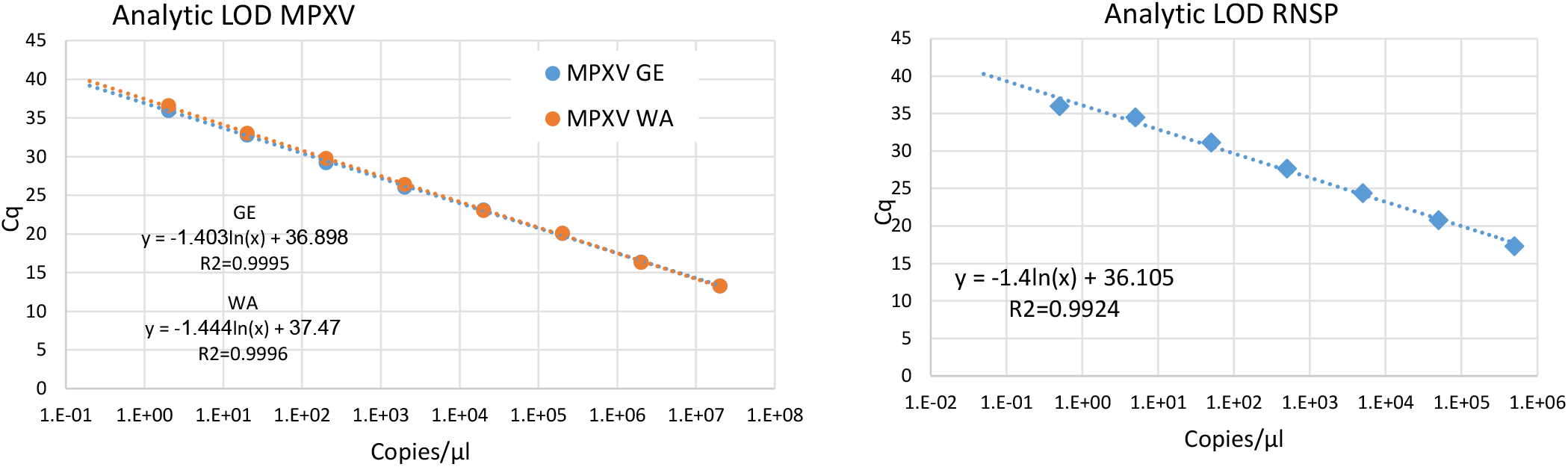
Analytic limit of detection (LOD) curves of standard quantified controls. Curves were plotted for the MPX GE and WA reactions (A) and for the human RNAse P reaction that was used as an endogenous control. Serial dilutions of quantified control DNA were tested in triplicates and the Cq values were plotted against the calculated concentration. The formula of the derived regression line and the R^2^ value of the curve are shown for each reaction.

